# Patterns of vascular access among chronic kidney disease patients on maintenance hemodialysis at Muhimbili National Hospital. A single centre cross-sectional study

**DOI:** 10.1101/2024.08.16.24312124

**Authors:** Daniel Msilanga, Jacqueline Shoo, Jonathan Mngumi

## Abstract

**Background:** Hemodialysis access profoundly impacts the quality of care for chronic kidney disease (CKD) patients worldwide, with arteriovenous fistulas (AVFs) preferred for superior outcomes. Despite global guidelines, Sub-Saharan Africa, including Tanzania, faces challenges, as it relies heavily on nontunneled central venous catheters (CVCs) due to accessibility and financial constraints. We aimed to describe the pattern of vascular access use among CKD patients on maintenance hemodialysis at Muhimbili National Hospital.

**Methods:** A cross-sectional study to describe the pattern of vascular access among patients with CKD on maintenance hemodialysis therapy. Descriptive statistics were used to summarize the baseline characteristics and patterns of vascular access. Our study received ethical clearance from the Muhimbili National Hospital Research Ethics Committee (Ref: MNH/IRB/VOL.1/2024/005). All consent forms were written and provided in English or Swahili.

**Results:** We analysed 200 study participants, with a mean age of 53.3 (14.5) years. Almost all participants initiated hemodialysis with nontunneled central venous catheters (95.5%). A substantial portion continued to use nontunneled CVCs (25.5%), with some transitioning to tunneled CVCs (39.5%) or AVFs (35%). The mean (SD) duration to use nontunneled CVCs before transitioning to tunneled or AVF were 7.1 (2.1) months. Among patients with multiple nontunneled catheters, catheter dislodgement was the main indication for catheter replacement.

**Conclusion:** Our study highlights the prevalent use of nontunneled central venous catheters (CVCs) as the primary vascular access method for CKD patients undergoing hemodialysis at Muhimbili National Hospital, Tanzania. These findings underscore the urgent need for analysis of the cost associated with nontunneled catheter reliance and interventions to improve access to AVFs and enhance vascular access management, ultimately optimizing patient outcomes in resource-limited settings.

## Introduction

Hemodialysis access is a critical component affecting the quality of care for chronic kidney disease (CKD) patients in need of renal replacement therapy (1,2). Worldwide, the Kidney Disease: Improving Global Outcomes (KDIGO) guidelines and the Fistula First Initiative advocate for arteriovenous fistulas (AVFs) as the preferred vascular access method due to their superior long-term outcomes and overall survival (3,4). Despite these recommendations, the problem of hemodialysis access remains substantial, with disparities observed in access patterns across different regions (5).

Adequate vascular access is pivotal for comprehensive care in chronic kidney disease (CKD) patients undergoing hemodialysis (1). Ideally, a fistula should be established six months prior to treatment initiation in suitable cases (2,6). Tunnelled catheters are reserved for patients with poor AVF outcomes due to vascular issues (7,8), while nontunnelled central venous catheters (CVCs) are discouraged for chronic use, mainly due to infection risks and vessel damage (9,10). Despite these recommendations, in sub-Saharan Africa, nontunneled CVCs are commonly used due to their accessibility and cost, especially in emergency situations where immediate dialysis is necessary, which often occurs out of pocket (1,11)

Like in many other parts of sub-Saharan Africa, hemodialysis is the mainstay kidney replacement therapy for most, while kidney transplantation remains a limited option in Tanzania (12). Patients undergoing maintenance hemodialysis in Tanzania face challenges accessing appropriate vascular access due to the limited availability of centers offering tunnelled dialysis central venous catheter (CVC) placement and arteriovenous fistula creation, compounded by financial constraints (5,11). As a result, many patients rely on nontunneled CVCs, which increases the risk of vascular complications and contributes to morbidity and mortality. This study aimed to describe the current practices and patterns of vascular access among CKD patients on maintenance hemodialysis, shedding light on current practices and identifying gaps in optimal vascular access management.

## Methodology

A cross-sectional study conducted at the Muhimbili National Hospital (MNH) hemodialysis units from 10^t^ April 2024 to 15^th^ May 2024. The MNH is a tertiary-level public health facility with a 1500-bed capacity. The MNH has 50 hemodialysis machines and dialyzes between 100 and 130 CKD patients per day.

### Recruitment procedure

During each dialysis visit, patients were identified from the dialysis appointment registry and randomly selected using a rotary method. The study details, including the study’s purpose, procedures, and potential risks and benefits, were explained to them, and they were invited to consent to participate in the study. Those who consented were included in the study. This process was repeated until the sample size was reached. Approximately half of the patients are covered by health insurance, whereas the remainder pay out-of-pocket for hemodialysis treatments. Due to financial constraints, almost all of those who are paying out-of-pocket are receiving dialysis therapy either twice or once weekly.

The data were collected through an interview-administered questionnaire. Selected clinical data, including comorbidities (diabetes/hypertension), frequency of hemodialysis treatments per week, and type of vascular access, were collected from the dialysis registry (a paper document). Vascular access type data included the type of initial access, anatomical location of vascular access, duration of access use, and current access, and to avoid the possibility of confusing dialysis access types, a physical examination of each patient was performed to ensure that proper vascular access was recorded. The outcome was the pattern of vascular access use during the course of dialysis therapy.

### Data analysis

escriptive statistics were used to summarize the baseline characteristics and patterns of vascular access.

Authors Access to Study Participant Information and Confidentiality: All information related to study participants will remain confidential and will be identifiable only by codes known to the researcher. To ensure participant privacy, all personal identifiers will be replaced with unique codes. Only the primary researcher will have access to the code key linking participants to their data. This process will safeguard participant confidentiality throughout the study

### Ethical approval

Ethical clearance was obtained from Muhimbili National Hospital, Clinical Research, Training and Consultancy Unit with reference number MNH/IRB/VOL.1/2024/005. All participants provided written informed consent before any study procedures are conducted. The consent form documents were written and provided in English and Swahili language.

## Results

Two hundred study participants were enrolled in this study. The mean age was 53.3 years (14.5), with almost half of the study participants being male (58.5%), unemployed (52.5%), under medical insurance (56%), and doing thrice weekly dialysis sessions (56.5%). Hypertension (46%) was the main etiology of CKD, followed by diabetes mellitus (30.5%). (Table 1)

**Table 1:** Demographic data (n=200)

|  | N (%) |
| --- | --- |
| Mean age (years) | 53.3 (14.5) |
| Gender |  |
| Male | 117 (58.5) |
| Female | 83 (41.5) |
| Occupation |  |
| Unemployed | 105 (52.5) |
| Retired | 48 (24.0) |
| Employed | 47 (23.5) |
| Payment modality for HD |  |
| Insured | 112 (56.0) |
| Uninsured (out-of-pocket payment) | 88 (44.0) |
| Etiology of CKD |  |
| Hypertension | 92 (46.0) |
| DM | 61 (30.5) |
| HIV | 15 (7.5) |
| Glomerulonephritis | 14 (7.0) |
| Unknown cause | 13 (6.5) |
| Other causes | 5 (2.5) |
| Weekly frequency of hemodialysis sessions |  |
| Three times per week | 113 (56.5) |
| Two times per week | 80 (40) |
| Once per week | 7 (3.5) |

CKD: Chronic kidney disease, DM: Diabetes mellitus, HD: Hemodialysis

Almost all study participants started hemodialysis therapy via a nontunneled central venous catheter (95.5%), and the rest had early arteriovenous fistulas (4.5%). Furthermore, a significant proportion of patients continued to rely on nontunneled central venous catheters as their access method (25.5%), while others transitioned to tunneled central venous catheters (39.5%) or arteriovenous fistulas (AVFs) (35%) (Table 2).

**Table 2:** Type of hemodialysis access (N=200)

|  |  |
| --- | --- |
|  | Paying from pocket (N=200) |

| Type of access | Initial access | Current access |
| --- | --- | --- |
| Nontunneled CVC | 191(95.5) | 51(25.5) |
| Tunneled CVC | 0 | 79(39.5) |
| AVF | 9 (4.5) | 70(35) |

CVC: Central venous catheter, AVF: arteriovenous fistula

Among the subset of prevalent ESRD patients who used only nontunneled CVCs for dialysis (25.5%), the mean duration of nontunneled CVC use was 7.1(2.1) months. A history of multiple nontunneled CVC catheterizations was reported in 68.6% of patients, and dislodgement of the catheter was the most common reason for recatheterization Table 3.

**Table 3:**
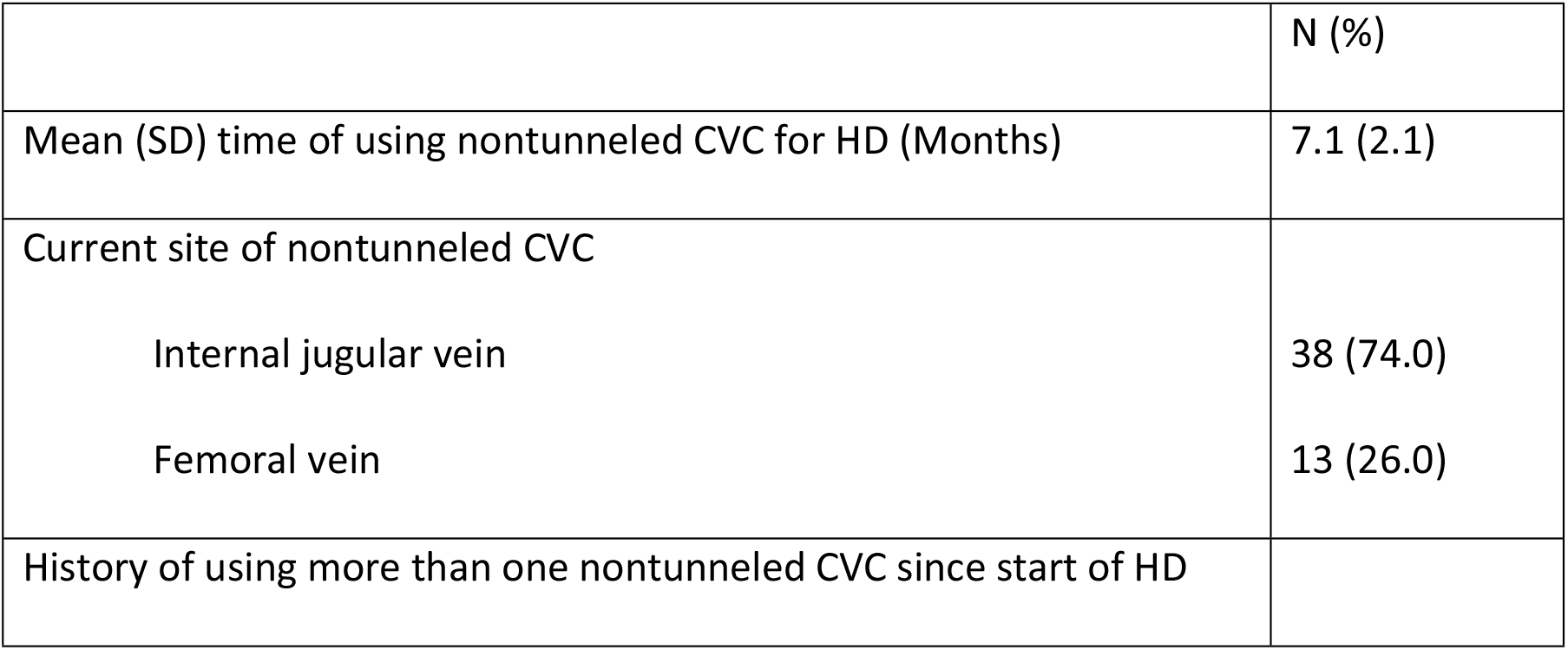
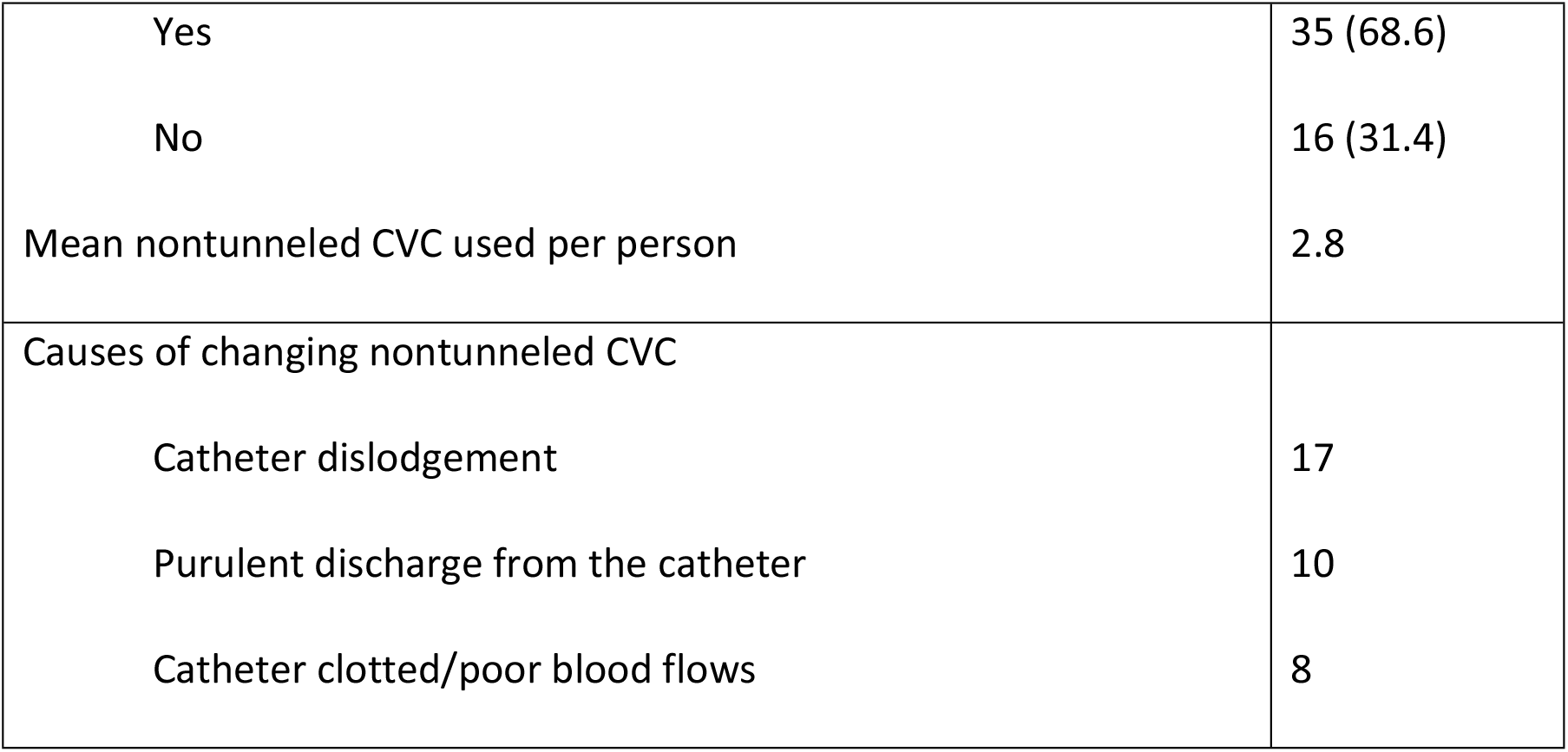
Patients with nontunneled central venous catheters as the only access for hemodialysis (N=51)

|  | N (%) |
| --- | --- |
| Mean (SD) time of using nontunneled CVC for HD (Months) | 7.1 (2.1) |
| Current site of nontunneled CVC |  |
| Internal jugular vein | 38 (74.0) |
| Femoral vein | 13 (26.0) |
| History of using more than one nontunneled CVC since start of HD |  |
| Yes | 35 (68.6) |
| No | 16 (31.4) |
| Mean nontunneled CVC used per person | 2.8 |
| Causes of changing nontunneled CVC |  |
| Catheter dislodgement | 17 |
| Purulent discharge from the catheter | 10 |
| Catheter clotted/poor blood flows | 8 |

## Discussion

Our study provides valuable insights into the status and vascular access practice patterns among patients with chronic kidney disease (CKD) undergoing maintenance hemodialysis therapy in Tanzania. Notably, nine out of ten study participants initiated hemodialysis therapy using nontunneled catheters. Subsequently, three-quarters of these participants transitioned to either tunnelled catheters or arteriovenous fistulas (AVFs), with a mean time to conversion from nontunnelled catheters of seven months. Additionally, a quarter of the participants continued to use nontunneled catheters for permanent dialysis access.

Vascular access serves as the lifeline for patients undergoing dialysis therapy (13,14). We found that almost all of the participants were initiated on hemodialysis therapy using nontunneled catheters. This trend mirrors findings reported in Kenya, Libya and Senegal, where more than 80% of incident CKD patients started hemodialysis therapy with a nontunneled catheter as the initial vascular access (15,16)(17). However, this is contrary to the practice in most higher income countries (HICs), where CKD patients start hemodialysis therapy either by using a tunnelled CVC and converting to an AVF or starting with an AVF (18–20). Despite the reported high rates of morbidity and mortality, nontunneled CVCs remain an important type of initial hemodialysis access for the majority of advanced CKD patients at the time of initiation of hemodialysis in LMICs. Likely contributors to this practice pattern include limited predialysis care, late presentation necessitating urgent hemodialysis and lack of expertise for tunneled CVCs or AVF (10,21).

Almost a quarter continued to rely on nontunneled catheters for permanent access, with a prolonged mean duration of 7.1 months. Similar findings were reported in Kenya and Senegal (15,16), but the mean duration of nontunneled catheter use was longer in our setting than the 3 months reported in other centers with the same economic settings (15,17). While nontunneled CVCs are employed as a temporary measure, their extended utilization raises major concerns regarding the associated risks, including infection, thrombosis, and mechanical complications leading to multiple admissions and high cost. A number of those who chronically utilized nontunneled CVCs had a history of multiple insertions. This highlights the challenges in maintaining vascular access with nontunneled catheters, increasing overall healthcare resource utilization, and increasing the cumulative risk of complications for patients.

Our study revealed the extensive reliance on nontunneled central venous catheters (CVCs) for hemodialysis among chronic kidney disease patients in Tanzania and similar settings with the same economic status and healthcare financing methods. This highlights the difficulty in accessing recommended vascular access options in resource-limited settings. The prolonged use of CVCs and frequent catheter insertions emphasize the need for improving access to arteriovenous fistulas (AVFs). Addressing these challenges is vital for minimizing infection and other complications and improving patient outcomes while reducing healthcare resource use. Targeted interventions and resource allocation are crucial for optimizing vascular access practices and improving care for hemodialysis patients in similar settings.

### Limitation

While we provided an overview of the vascular access patterns among CKD patients on maintenance hemodialysis at MNH, we could not analyse potential factors influencing these patterns or their outcomes. Despite this limitation, our study sheds light on the significant burden of relying on nontunneled central venous catheters (CVCs) for hemodialysis in LMICs, such as Tanzania. In addition, our findings emphasize the need for further research to assess the costs associated with the prolonged use of nontunneled catheters and its complications.

## Data Availability

All data will be available and accessible upon request to the authors

## Data availability statement

The datasets generated and/or analysed during the current study are available from the corresponding author on reasonable request. All data supporting the findings of this study are included within the article

## Acknowledgements

We thank the Muhimbili National Hospital for approving the research and extend our deep gratitude to the patients who participated in the study.

